# Which electronic health record system should we use? – a systematic review

**DOI:** 10.1101/2020.10.11.20210930

**Authors:** Mohammed Al Ani, George Garas, James Hollingshead, Drostan Cheetham, Thanos Athanasiou, Vanash Patel

## Abstract

**Objectives:** This is the first systematic review to look at all published data on EHRs to determine which systems are advantageous.

**Design:** A systematic review was performed by searching EMBASE and Ovid MEDLINE between 1974 and November 2019.

**Participants:** All original studies that appraised EHR systems were included.

**Main outcome measures:** EHR system comparison, implementation, user satisfaction, efficiency and performance, documentation, and research and development.

**Results:** The search strategy identified 701 studies, which were filtered down to 46 relevant studies. Level of evidence ranged from 1 to 4 according to the Oxford Centre for Evidence-based Medicine. The majority of the studies were performed in the USA (n = 44). N=6 studies compared more than one EHR, and Epic followed by Cerner were the most favourable through direct comparison. N=17 studies evaluated implementation which highlighted that it was challenging, and productivity dipped in the early phase. N=5 studies reflected on user satisfaction, with women demonstrating higher satisfaction than men. Efficiency and performance issues were the driving force behind user dissatisfaction. N=26 studies addressed efficiency and performance, which improved with long-term use and familiarity. N=18 studies considered documentation and showed that EHRs had a positive impact with basic and speciality tasks. N=29 studies assessed research and development which revealed vast capabilities and positive implications.

**Conclusion:** Epic is the most studied EHR system and the most commonly used vendor on the market. There is limited comparative data between EHR vendors, so it is difficult to assess which is the most advantageous system.

## INTRODUCTION

An Electronic Health Record (EHR) is an electronic version of a patients medical history, that is maintained by the provider over time, and may include all of the key administrative clinical data relevant to that person’s care.[1] Several sectors of the UK’s health system use EHRs to a varying degree. These include National Health Service (NHS) hospitals, primary care (general practice) and social care services. In the UK, the NHS 5-year forward plan outlined its desire to cut the use of paper records and go fully digital. NHS England intends to connect EHRs across primary, secondary and social care by 2020.[2] There are several challenges to implementing EHR systems across the NHS. These include interoperability of information technology systems, system installation and staff training, opportunities for patient access, consequences for the Doctor-Patient relationship, and data security and privacy.[3] Currently, the majority of software for Summary Care Records (SCR), which is an electronic record of important patient information, are created for General Practice (GP) medical records. They can be seen and used by authorised staff in other areas of the health and social care system involved in the patient’s direct care. Egton Medical Information Systems (EMIS), TPP and inPractice provide SCRs. Large suppliers of hospital software include Cerner, Computer Sciences Corporation (CSC), British Telecom (BT) and IMS Maxims. Open Source software, which is software with source code that anyone can inspect, modify, and enhance is also increasingly used, and has been endorsed by NHS England.[3] The objective of this review is to identify studies that compare EHRs; in terms of direct comparison between systems, implementation, documentation, user satisfactions, performance/efficiency and, research and development.

## METHODS

This study was performed following guidelines from the Preferred Reporting Items for Systematic reviews and Meta-Analyses (PRISMA).[4]

### Data sources and searches

Studies to be included in the review were identified by searching the following databases: (1) EMBASE (1974–November 2019) and (2) Ovid MEDLINE (1974–November 2019). All databases were searched using the following free text search: ‘electronic health records OR electronic medical records OR electronic Care Record OR NHS Care Records Service’ AND ‘Allscripts OR Cerner OR DXC OR IMS OR Maxims OR Nervecentre OR Meditech OR System C OR Elation OR ChartLogic OR Sevocity OR InSync Healthcare Solutions OR PrognoCIS OR Kareo OR EMA OR Epic OR EncounterWorks OR Azalea OR MicroMD OR ReLiMed OR Practice Fusion OR CareCloud Charts OR MedEZ OR AdvancedEHR OR All in One EHR OR Prime Clinical OR InteGreat OR Clarity EHR OR 1st Providers Choice EHR OR EyeMD EMR OR WRS Health EMR OR Aprima Medical Software OR TRAKnet ORPraxis EMR OR WebPT EMR OR OncologyCloud OR Centricity EMR OR eClinicalWorks 10e OR Practice Choice OR Medisoft Clinical OR Lytec MD OR Practice Partner OR athenahealth OR TherapyAppointment OR EchoVantage OR OmniMD OR RXNT OR AZZLY rize OR Intergy OR Zoobook OR ClinicTracker OR Nexus EHR OR Care360 OR Quanum OR CentralReach OR Blue EHR OR BestNotes OR MDVision OR Office Practicum OR NextGen Healthcare OR PulseEHR OR RevolutionEHR OR mds:chart OR DocuTAP OR VitalHealth EHR OR Allergy EHR OR Exscribe OR ICANotes OR Remedy OR Carelogic OR drchrono OR VelociDoc OR ECLIPSE OR ChiroTouch OR ReDoc OR AgilityOM OR PIMSY OR PT Practice Pro OR Renesan OR TSI OR Valant OR Sleep WorkFlow OR TherapyNotes OR Falcon Physician OR AllMeds OR eDerm OR Medios OR Amazing Charts EHR OR IMS Clinical OR ClinicSource OR FreePT OR OfficeEMR OR Medflow OR NueMD OR SOAPware OR iPatientCare OR PrimeSuite’.

### Study selection

We included all original studies that compared more than one EHR system. We also included any study that looked at various aspects pertaining to EHR systems. These included studies that looked at implementation, user satisfaction, efficiency/performance, documentation, and research and development. Studies that mentioned EHR or any of the search criteria as a tool for data collection only were excluded. Abstract studies were excluded. Two authors (MA and VP) independently reviewed the titles and abstracts of the retrieved articles, and selected publications to be included in this review. The full texts of these publications were reviewed by the two authors, who then selected the relevant articles for inclusion in the review.

### Data extraction and quality assessment

Two authors (MA and VP) independently extracted data from the full text, which included source of article, study design, study period, type of EHR System, system comparison, implementation, user satisfaction, efficiency and performance, documentation, and research and development. Study design and study quality was determined using the Oxford Centre for Evidence-Based Medicine (EBM) study design and levels of evidence classification, respectively.[5, 6]

### Data synthesis and analysis

The methodology of the included studies was heterogeneous; therefore, it was not possible to pool data to conduct statistical analysis.

## RESULTS

### Study selection

701 potentially relevant articles were retrieved. Of these, 600 articles were excluded following title and abstract review. Of the remaining 101 articles, 54 studies were excluded because they were abstracts only. Of the remaining 47 studies one paper was excluded because the full paper was not in the English language. The remaining 46 studies were included in this review (**Figure 1**) (**Table 1**). The agreement for inclusion of the studies between the authors was satisfactory (κ = 0.89).

**Table 1:**
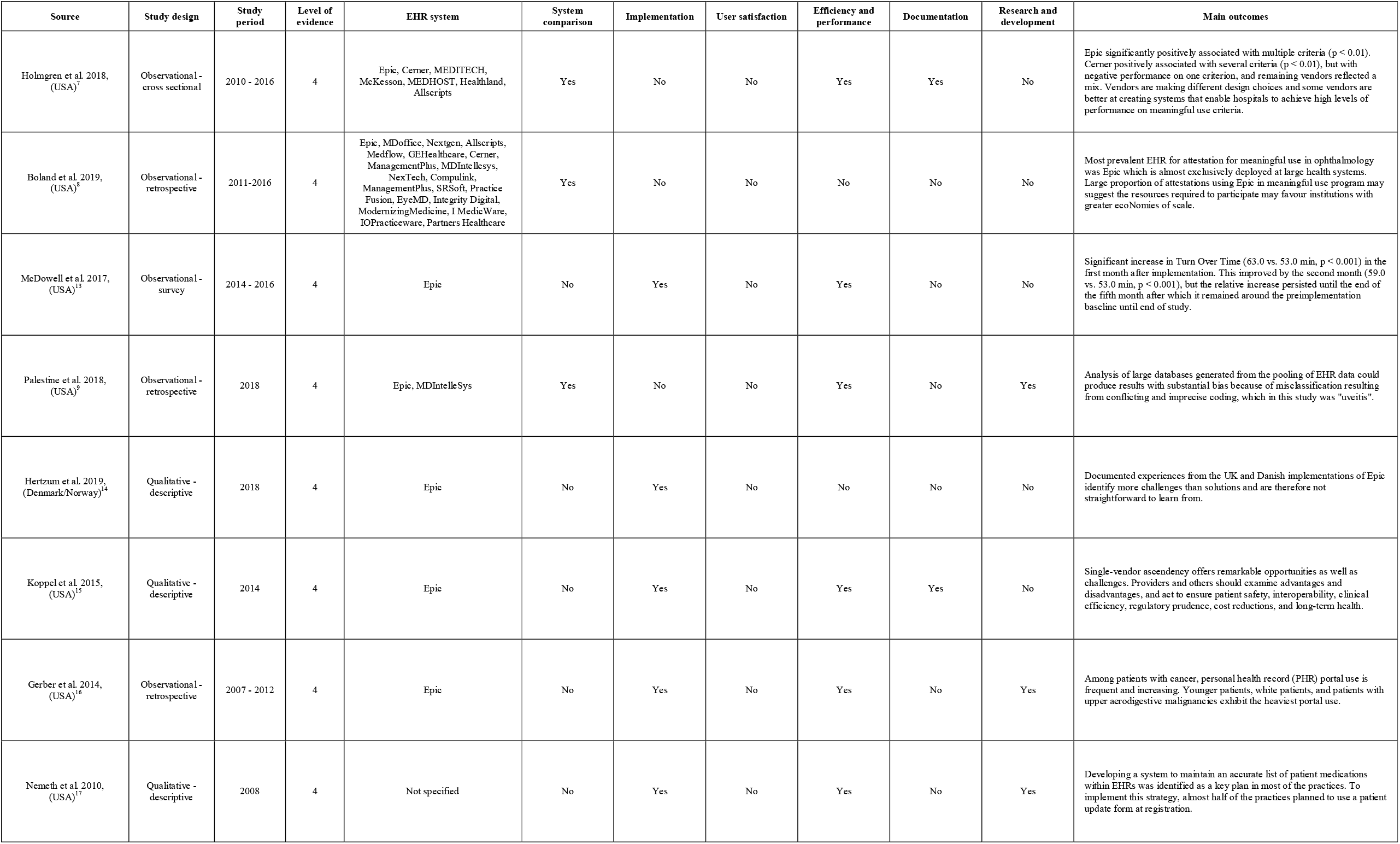
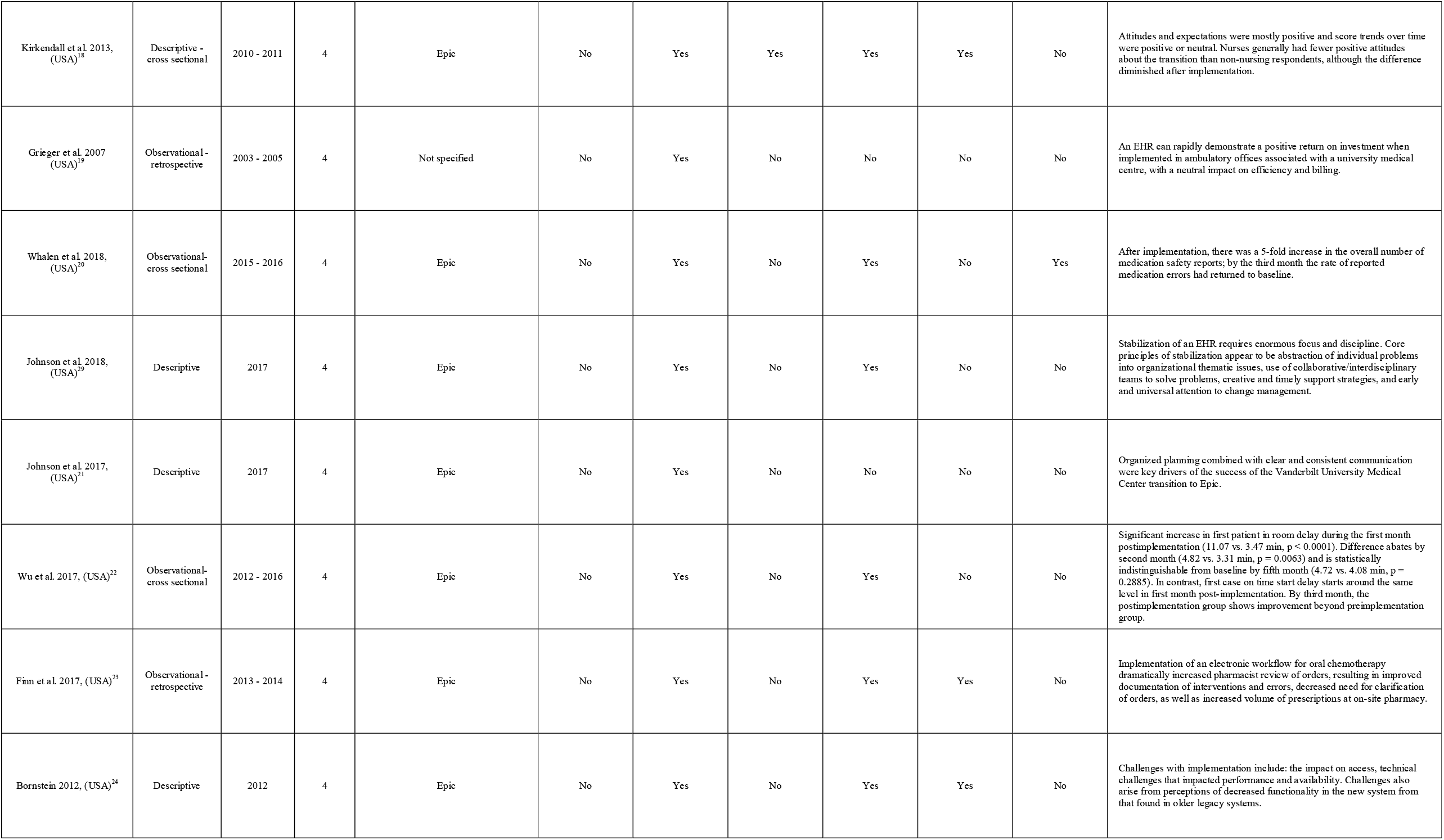
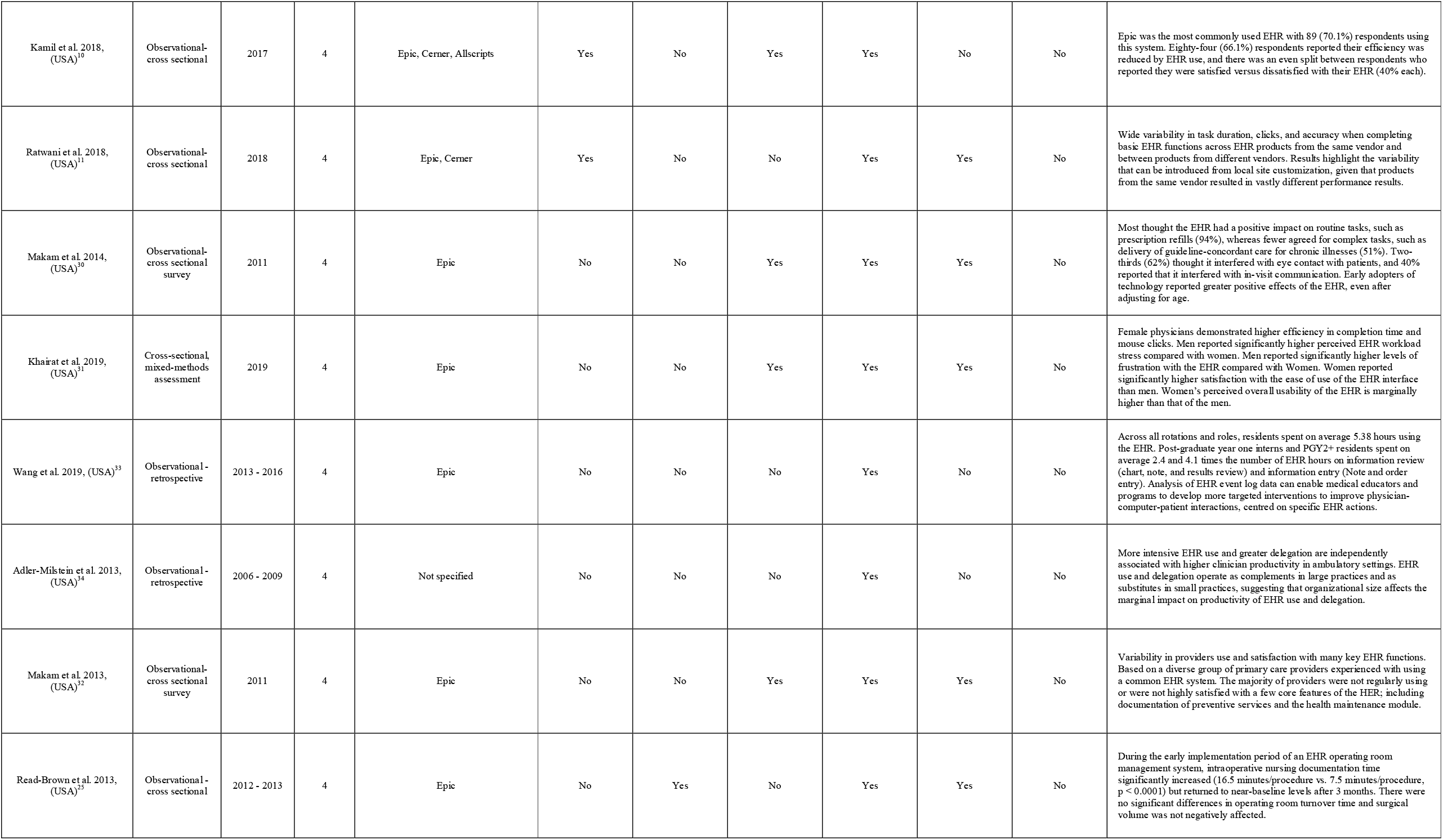
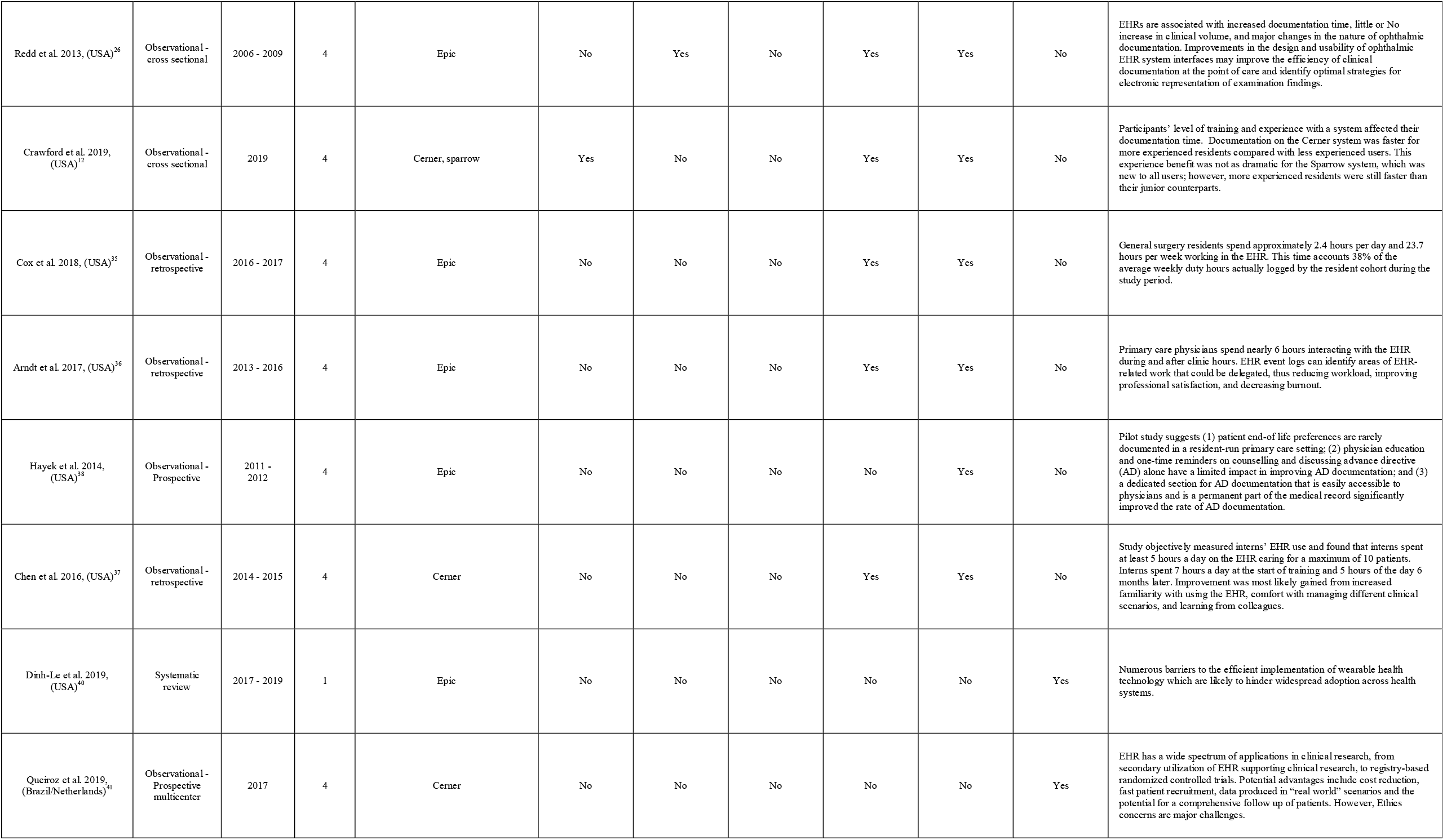
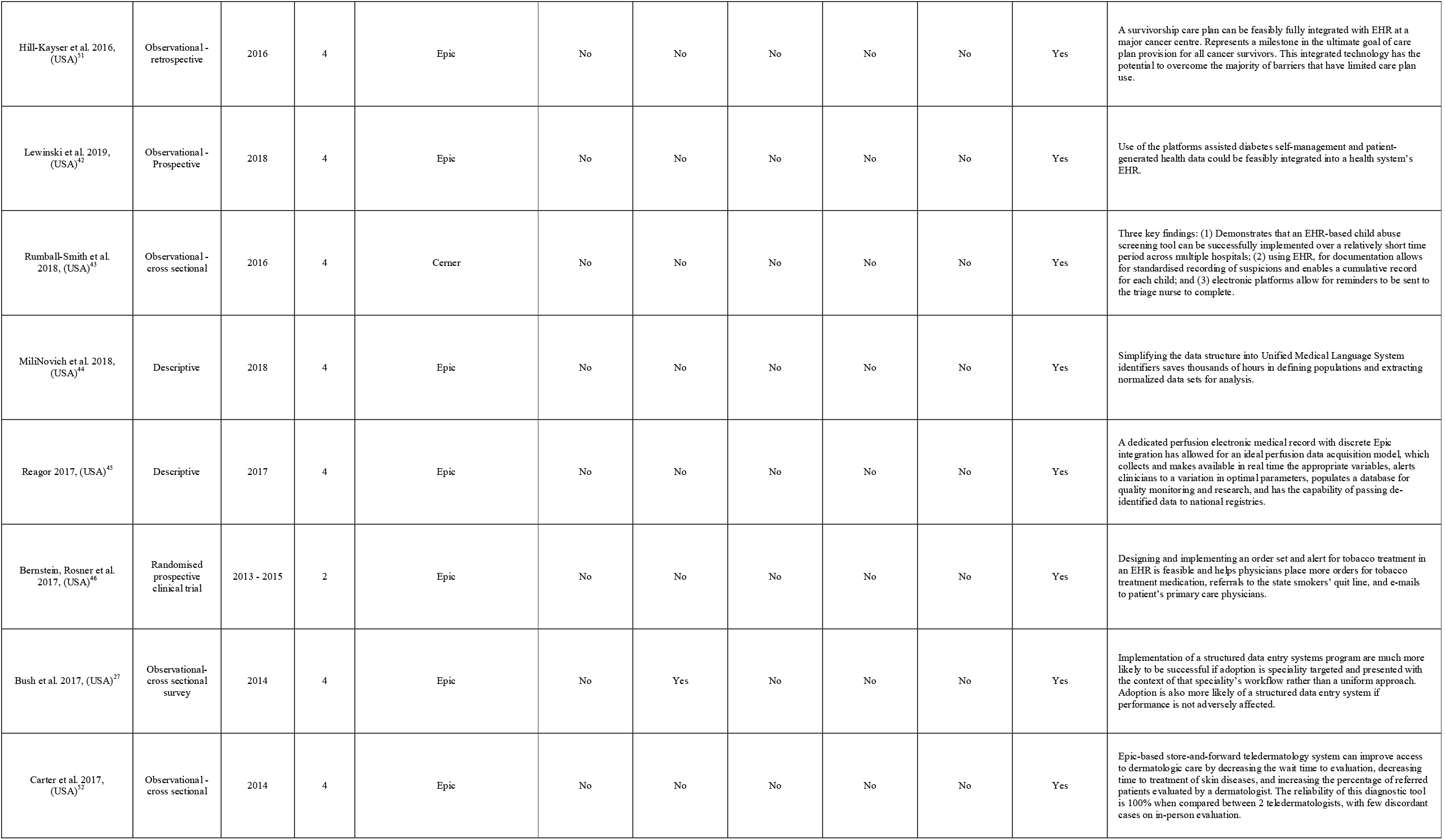
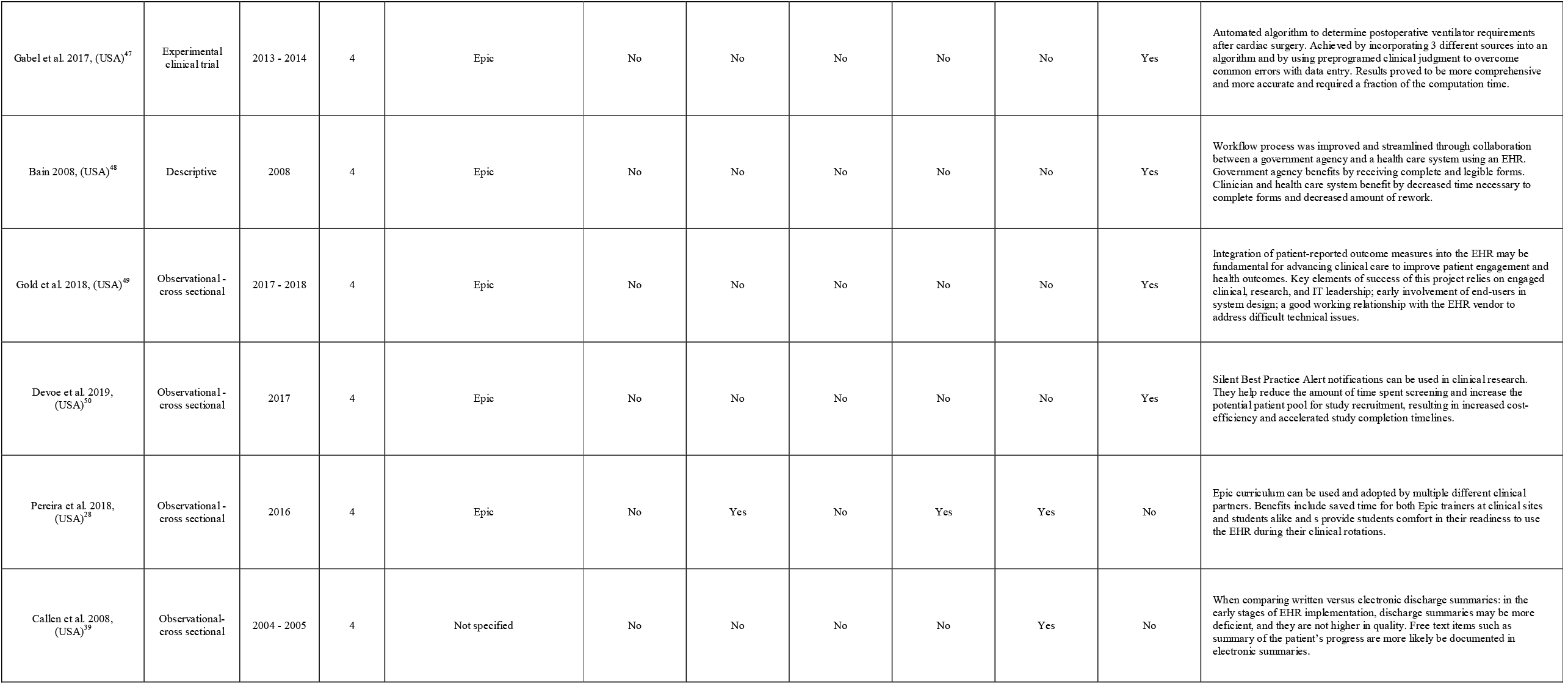
Description of studies included in the systematic review

**Figure 1:**
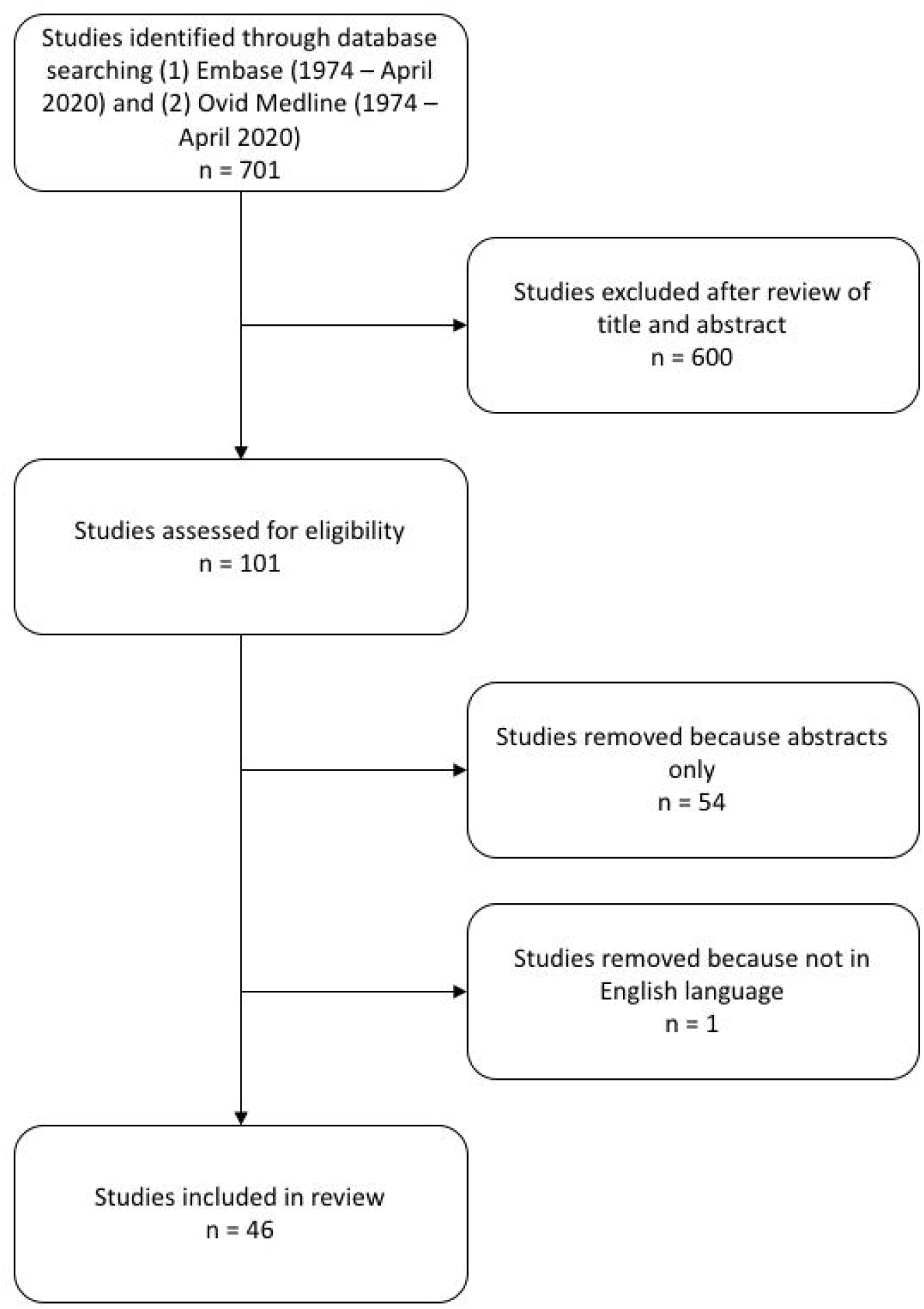
Selection of articles for the systematic review

### Study characteristics

The level of evidence ranged from levels 1 to 4 according to the Oxford Centre for Evidence-based Medicine (**Table 1**).[6] The bulk of the studies were performed in the USA (n = 44), one study represented a collaborative between Brazil and the Netherlands (n = 1) and the other was a collaborative between Denmark and Norway (n = 1). The studies were published from 2007 until 2019, with the majority published in the last 3 years (i.e., between 2017 and 2019, n = 30).

### System comparison

The following studies compared more than one EHR system in their study (n = 6). One study compared; Epic/ Cerner/ MEDITECH/ McKesson/ MEDHOST/ Healthland/ Allscripts/ Other.[7] Another study looked at Epic/Epic/MDoffice/Nextgen/ Allscripts/ Medflow/ GEHealthcare/ Cerner/ ManagementPlus/ MDIntellesys/ NexTech/ Compulink/ ManagementPlus/ SRSoft/ Practice Fusion/ EyeMD/ Integrity Digital/ ModernizingMedicine/ I MedicWare/ IOPracticeware/ Partners Healthcare.[8] There were also comparisons between Epic / MDIntelleSys [9], Epic / Cerner /Allscripts [10], Epic/ Cerner [11] and Cerner/ sparrow.[12] Epic was the most favourable system when it came to direct comparison with other EHR systems. Cerner was in second contention; however, this data was limited in terms of number of studies and range of system functions compared. Based on the study by Holmgren et al. 2018, (USA), Epic was associated with significantly better performance on 5 of the 6 criteria.[7] These criteria included the following; medication orders entered using computerized provider order entry, patient’s ability to View/Download/Transmit (VDT) their health information, VDT used, medication reconciliation, summary of care record provided and summary of care record sent electronically. In comparison Cerner was significantly positively associated with 3 of these criteria.

### Implementation

The following studies looked at various aspects of implementing an EHR system and its implications (n = 17).[13-28] There was a general consensus among the studies that implementation of EHR systems was challenging in the first instance. It required intense planning and support prior to and during the implementation phase. Productivity also dipped in the early phase of implementation, but several studies showed that productivity improved soon after to return to baseline or make further gains. McDowell et al. 2017, (USA) showed that operating room efficiency, as measured by turnover time of patients, decreased in the first five months post-implementation returning to baseline at six months.[13] Wu et al. 2017, (USA) showed that there was a significant increase in first person in Operating Room (OR) delay during the first month post-implementation.[22] This difference abated by the second month and becomes statistically indistinguishable from baseline by the fifth month. Read-Brown et al. 2013, (USA) showed a similar dip in performance for a similar duration of time.[25] It reported that intraoperative nursing documentation time significantly increased but returned to near-baseline levels within 3 months. Johnson et al. 2018, (USA) reported that it took 30 months of planning to go live to an epic platform where 17,000 employees were affected by the switch.[29] A study that looked at the UK and Denmark’s implementation experience reported that it took Cambridge University Hospitals NHS Foundation Trust 18 months from signing its contract with Epic to going live,[14] while in Denmark where the contract with Epic was signed in 2013, the first implementation began in 2016 with the last of the hospitals making the switch in 2017.

### User satisfaction

Of the 5 studies (n=5) that evaluated user satisfaction, they all related to Epic [10, 18, 30-32] except for one that also examined Cerner and Allscripts.[10] There were mixed reviews when it came to user satisfaction. Issues with efficiency and performance was the driving force behind user dissatisfaction with EHR usage overall. Kamil et al. 2018, (USA) showed that only 37.8% (n = 48) of respondents reported they were somewhat or very satisfied with the EHR while 40.9% (n = 52) of respondents reported they were somewhat or very dissatisfied with the EHR.[10] The five most frequently cited challenges were excessive time burden (n = 36, 28.3%), increased work associated with regulatory compliance (n = 17, 13.4%), the cumbersome, poor design of the EHR (n = 16, 12.6%), the lack of neurotology-specific customization of the EHR (n = 15, 11.8%), and the focus away from patients (n = 14, 9.55%). Makam et al. 2014, (USA) reported that providers expressed concerns about the impact of the EHR on in-person communication with patients.[32] Nearly two-thirds (62%) agreed that the EHR made it difficult to maintain eye contact with their patients, and 40% thought it interfered with provider–patient communication during visits. There was also an interesting gender difference when it came to EHR user satisfaction. The study by Khairat et al. 2019, (USA)[31] showed that men had a higher perceived EHR workload stress compared with women (p < 0.001). Men reported significantly higher levels of frustration with the EHR compared with women (p < 0.001). Women reported significantly higher satisfaction with the ease of use of the EHR interface than men (p = 0.03), and women’s perceived overall usability of the EHR is marginally higher than that of the men (p = 0.06).

### Efficiency and performance

EHR efficiency and performance was the most studied aspect (n =26).[7, 9-13, 15-18, 20-26, 31-37] Long-term use and familiarity with the EHR system helped to improve efficiency. However, the overall review was poor in terms of efficiency and performance with the most commonly cited issues relating to the time taken to complete tasks. This was particularly challenging for those new to EHR and required extra time outside of normal working hours spent on completing various tasks. Kamil et al. 2018, (USA) showed that a majority of responders (n = 84, 66.1%) reported significantly or mildly decreased efficiency.[10]

Makam et al. 2014, (USA) showed that each 5-year decrease in the number of years since graduation was associated with a 16% increase in the odds of agreeing that EHR was easy to use; a 15% increase in the odds of agreeing the EHR was easy to learn; a 25% increase in the odds of agreeing the EHR improved clinical workflow; a 27% increase in the odds of agreeing that they like to discover new ways to use the EHR; and a 12% increase in the odds of agreeing that EHR helps them be more thorough.[30]

### Documentation

The following studies looked at documentation using EHR (n=18).[7, 11, 12, 15, 18, 23-26, 30, 31, 35-39] Studies showed that documentation with EHR had a positive impact, especially with basic tasks. Its improved productivity, when tailored to specific speciality requirement, allowed a more streamline experience. It also allowed for errors to be picked up and addressed in a more effective manner. Finn et al. 2017, (USA) showed that implementation of Epic Beacon for documentation allowed for identification of over 500% more chemotherapy order errors (41 total errors identified pre-Beacon versus 250 total errors identified post-Beacon).[23] Makam, et al. 2014, (USA) showed that most thought the EHR had a positive impact on routine tasks, such as prescription refills (94%), whereas fewer agreed when it came to complex tasks, such as the delivery of guideline-concordant care for chronic illnesses (51%).[30]

### Research and development

The following studies looked at using EHR in research (n=14)[9, 16, 27, 40-50] and development (n= 15).[20, 28, 31, 38, 40-43, 45, 47-52] Several studies looking at EHR in research showed its vast capabilities and positive implications. There is huge potential to using EHR for collecting large volume of data efficiently and streamline patient information at the port of entry to alert for specific concerns. Development with third party software and gadgets is also being looked at, although this is an emerging market that should allow patients more freedom to review and input their health data. Hayek et al. 2014, (USA) implemented a reminder system consisting of the addition of ‘‘Advanced Directives Counselling’’ to the problem list of EHR, and they managed to increase advance directive documentation to 76% compared to 11.5% without the reminder prompting.[38] Queiroz et al. 2019, (Brazil/Netherlands) used automatization of data collection for clinical research using EHR and demonstrated that 76.5% to 100% of accurate data can be obtained from this setup.[41]

## DISCUSSION

This is the first systematic review to compare different EHR systems. What becomes evident is there are limited studies conducted outside of the USA on the subject, where the leading market share is held by Epic. Epic was the most studied EHR system and studies pertaining to Epic have looked at all aspects of the system from implementation to research and development. Switching to an EHR system is no easy venture, a huge amount of planning and recourses are required. An overall dip in performance is expected to come with the immediate transition. Intensive support across the board especially from the information technology teams is crucial. Documentation with EHR improves efficiency however becoming proficient at using EHR has its drawbacks and limitations. Certain EHR systems (Epic followed by Cerner) are better at this than others, although the numbers of studies looking at direct system comparisons are limited. EHR use in research has a lot of potential and development of software to enable patient-centred care is on the horizon with improvement in technology.

There is little doubt that a gold standard EHR system would be challenging to achieve as evident by the efficiency and performance studies. There are several different user groups (doctors, nurses, ward clerks, administrators, etc.), in different settings (Accident and Emergency, speciality wards, operating theatres, outpatient clinics, etc.) using the same system. However, an ideal system (**Figure 2**) should have, maximum IT support pre- and during its implementation phase. Continued training should be offered and along with a feedback platform for improvements. Training should start early at either medical school or nursing school and implemented into teaching syllabuses. This will allow for more effective transition into working life for healthcare professionals. Routine tasks such as requesting scans should not be taxing or complicated with multiple steps. Flexibility in the layout and templates to allow for diversity of use is essential across the different settings. Automatic populating of data should be offered to help expedite tasks sue as discharge summaries or on templates to be used when alerting against potential adverse events.

**Figure 2:**
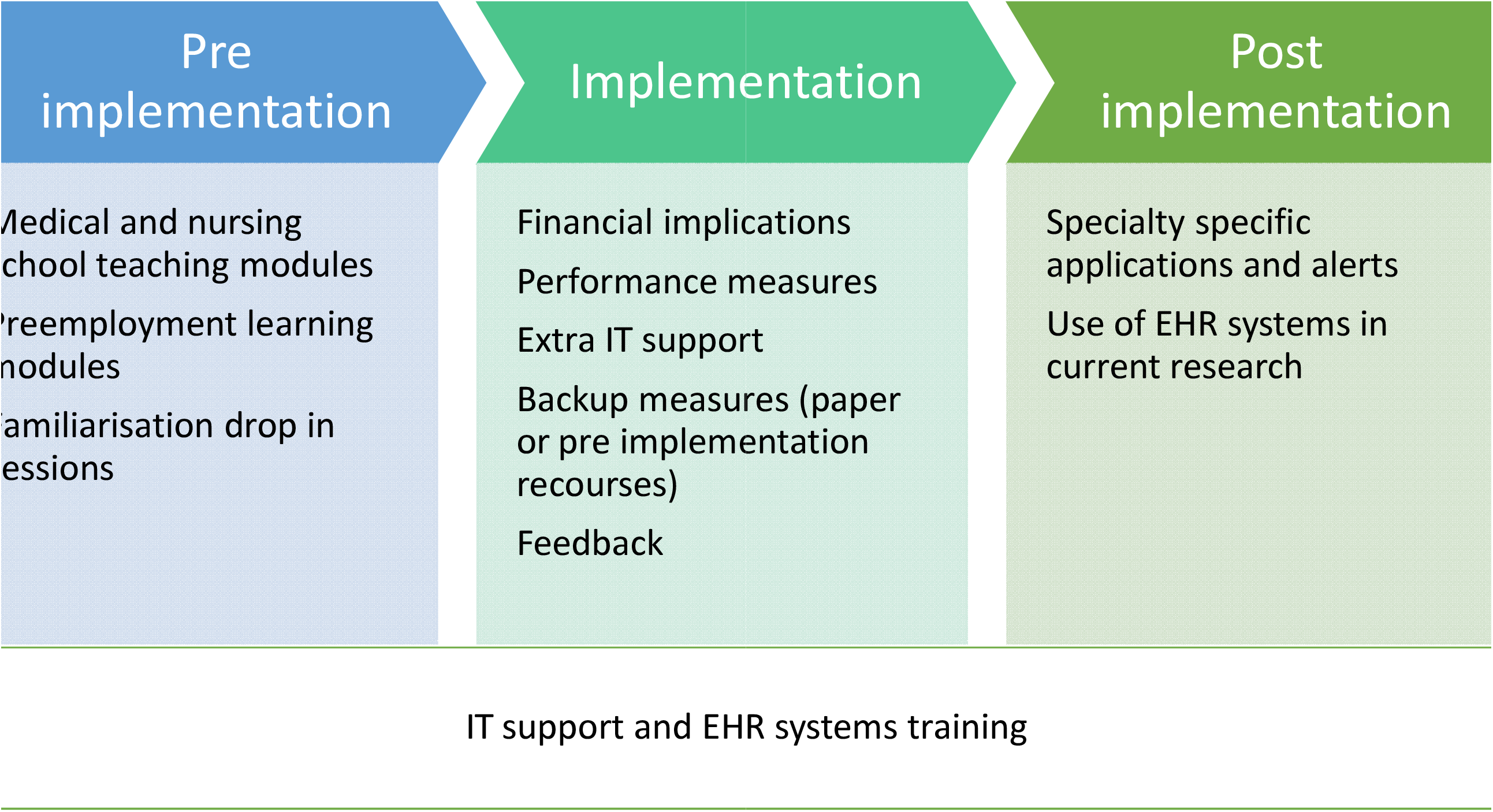
Process of implementing and EHR system

In terms of the strengths of this study, most of the studies were conducted in well-respected and known centres in the USA. They are also very current, and most were published in the last 3 years.

Most of the results of the studies came from one EHR system Epic, which can be seen as one of the limitations of this study. In addition, meta-analysis could not be performed due to the heterogeneity of the studies. This study clearly highlights the lack of comparison between EHR systems and the monopoly that Epic may hold across the USA. There was very limited data with regard to cost implications, which is a very important matter in today’s NHS. Other relevant aspects such as medicolegal implications, were barely touched on by one study in its survey findings, but not elaborated on this any further.[10] While effectiveness in clinical governance was not considered by any of the studies. More research and studies are required from UK-based hospitals on systems commonly used in the UK.

## CONCLUSION

Epic was the most studied EHR system and the most commonly used vendor on the market. There is limited comparative data between EHR systems and therefore it is difficult to assess which is the most advantageous system currently available.

## Data Availability

All data generated or analysed during this study are included in this published article (and its supplementary information files).

## SUMMARY POINTS

Already known:

- Epic is a popular and widely adopted EHR system.
- Challenges exist surrounding implementation of EHR systems, such as decreased productivity while operators adapt to a new system.
- Superiority of documentation of simple tasks using EHR versus paper systems.

What the study adds:

- Comparison of performance of several EHR systems.
- Deeper understanding of the lag period associated with implementation of EHR and productivity.
- Identifies gender discrepancy of user satisfaction with using an EHR system.
- Knowledge of how to use EHR in research and development.

## AUTHORS’ CONTRIBUTIONS

Mohammed Al Ani - literature search, study design, data collection, data analysis, data interpretation

George Garas – data analysis, data interpretation, critical revision

James Hollingshead – study concept, critical revision

Drostan Cheetham – study concept, critical revision

Thanos Athanasiou - study design, critical revision

Vanash Patel (corresponding author) – study design, data analysis, data interpretation, critical revision

All authors contributed to drafting the article or revising it critically for important intellectual content. All authors have approved the final version of the manuscript.

All authors had full access to all of the data (including statistical reports and tables) in the study and take responsibility for the integrity of the data and the accuracy of the data analysis.

## TRANSPARENCY DECLARATION

The corresponding author affirms that the manuscript is an honest, accurate, and transparent account of the study being reported. No important aspects of the study have been omitted, and any discrepancies from the study as planned have been explained.

